# Identification of a sex-specific genetic signature in dementia with Lewy bodies: a meta-analysis of genome-wide association studies

**DOI:** 10.1101/2022.11.22.22282597

**Authors:** Elizabeth Gibbons, Arvid Rongve, Itziar de Rojas, Alexey Shadrin, Kaitlyn Westra, Allison Baumgartner, Levi Rosendall, Zachary Madaj, Dena G. Hernandez, Owen A. Ross, Valentina Escott-Price, Claire Shepherd, Laura Parkkinen, Sonja W. Scholz, Juan C. Troncoso, Olga Pletnikova, Ted Dawson, Liana Rosenthal, Olaf Ansorge, Jordi Clarimon, Alberto Lleo, Estrella Morenas-Rodriguez, Lorraine Clark, Lawrence S Honig, Karen Marder, Afina Lemstra, Ekaterina Rogaeva, Peter St. George-Hyslop, Elisabet Londos, Henrik Zetterberg, Kevin Morgan, Claire Troakes, Safa Al-Sarraj, Tammaryn Lashley, Janice Holton, Yaroslau Compta, Vivianna Van Deerlin, Geidy E Serrano, Thomas G Beach, Suzanne Lesage, Douglas Galasko, Eliezer Masliah, Isabel Santana, Pau Pastor, Monica Diez-Fairen, Miquel Aguilar, Marta Marquie, Pablo Garcia-Gonzalez, Claudia Olive, Raquel Puerta, Amanda Cano, Oscar Sotolongo-Grau, Sergi Valero, Vanesa Veronica Pytel, Maitee Rosende-Roca, Montserrat Alegret, Lluis Tarraga, Merce Boada, Angel Carracedo, Emilio Franco-Macias, Jordi Perez-Tur, Jose Luis Royo, Jose Maria Garcia-Alberca, Luis Miguel Real, Maria Eugenia Saez, Maria Jesus Bullido, Miguel Calero, Miguel Medina, Pablo Mir, Pascual Sanchez-Juan, Victoria Alvarez, Kayenat Parveen, Kumar Parijat Tripathi, Stefanie Heilmann-Heimbach, Alfredo Ramirez, Pentti J. Tienari, Olivier Bousiges, Frederic Blanc, Chiara Fenoglio, Alessandro Padovani, Barbara Borroni, Andrea Pilotto, Flavio Nobili, Ingvild Saltvedt, Tormod Fladby, Geir Selbaek, Ingunn Bosnes, Geir Brathen, Annette Hartmann, Afina W. Lemstra, Dan Rujescu, Brit Mollenhauer, Byron Creese, Marie-Christine Chartier-Harlin, Lavinia Athanasiu, Srdjan Djurovic, Leonidas Chouliaras, John T. OBrien, Liisa Myllykangas, Minna Oinas, Tamas Revesz, Andrew Lees, Brad F Boeve, Ronald C. Petersen, Tanis J Ferman, Neill Graff-Radford, Nigel J. Cairns, John C. Morris, Glenda M. Halliday, John Hardy, Dennis W. Dickson, Andrew Singleton, David J. Stone, Ole A. Andreassen, Agustin Ruiz, Dag Aarsland, Rita Guerreiro, Jose Bras

## Abstract

**Background:** Genome-wide Association Studies (GWAS) have reshaped our understanding of the genetic bases of complex diseases in general and neurodegenerative diseases in particular. Despite being a common disorder, dementia with Lewy bodies (DLB), which, together with Parkinson’s disease dementia (PDD), comprise the umbrella term Lewy body dementias (LBD), is far from being well-characterized genetically. This is primarily due to a lack of familial cases and difficulty recruiting large, deeply characterized cohorts, given the high rate of misdiagnosis. By performing the largest GWAS in DLB, we aimed to identify novel risk loci to gain a better understanding of this disease’s pathobiology.

**Methods:** Here, we conducted the largest meta-analysis of genome-wide association studies performed in LBD, using a total of 5,119 cases and 20,988 controls, from five independent datasets, aggregating all previously published DLB genome-wide association results to date, as well as two previously undescribed cohorts. Additionally, we performed a sex stratified GWAS using the discovery datasets. We updated the heritability estimates for DLB and, to fine map these estimates, we used local heritability analysis. We calculated genetic correlation estimates between DLB and a range of other diseases and traits to identify potential pleiotropy. We also performed gene-set analysis to identify genes with excess burden of rare variability and pathway analysis. Lastly, we used the UK Biobank data to perform a PheWas using individuals at the extremes of genetic risk for DLB.

**Findings:** Between November 2018 and September 2022 we analyzed 8.6 million single nucleotide polymorphisms in 3293 DLB cases, 1826 LBD cases and 20,988 controls, as well as phenotypes from the UK Biobank dataset. Despite more than doubling the sample size from the previous GWAS in DLB, we did not identify significant loci in addition to those previously reported at *GBA, SNCA, STX1B*, and *APOE*. However, the sex-stratified analysis revealed that the *GBA* and *SNCA* signals are mainly driven by males, suggesting a sex-specific genetic architecture of disease. Using only clinical and neuropathologically diagnosed cases, we highlight four loci surpassing the significance threshold. Using the largest cohort of DLB we update our heritability estimates to 13% and fine map these results highlighting regions of the genome with high heritability but no genome-wide significant result so far.

**Interpretation:** These data provide the most comprehensive analysis of genetic variability in DLB to date. The fact that no novel risk loci have been identified after doubling the cohort size indicates the potentially significant role of rare variants in the genetic architecture of DLB and stresses the urgent need for larger, well-characterized cohorts of this disease for genetic studies. The sex-stratified analysis shows that males and females have different signatures of genetic risk for DLB. These results have widespread implications for clinical practice and clinical trials’ design in DLB.

## Introduction

Lewy body dementia (LBD) is an umbrella term encompassing two clinically distinct disease entities: dementia with Lewy bodies (DLB) and Parkinson’s disease dementia (PDD). DLB is the second most common form of dementia after Alzheimer’s disease (AD) and comprises up to 30% of all dementia cases ^1^. It is characterized by progressive cognitive decline and additional symptoms, such as visual hallucinations, parkinsonism, and rapid eye movement sleep behavior disorder ^2^. Despite this prevalence, the disease remains highly understudied ^3^. This disease’s clinical and pathological hallmarks make diagnosing it challenging, as they strongly overlap with Alzheimer’s and Parkinson’s diseases (PD) ^4^. It is currently estimated that approximately 20% of DLB diagnoses are incorrect ^5^. In addition, the only reliably replicated genetic signals involved in DLB (*SNCA, GBA*, and *APOE)* are also prominent features of the genetic architecture of both AD and PD ^6,7^, and therefore lack specificity for DLB.

Recent work has focused on analyzing the genetics of DLB in large cohorts to identify novel associations. In 2018, the first DLB GWAS was conducted on 1,743 cases and 4,454 controls, detecting significant associations at *GBA, SNCA, APOE* and other suggestive signals ^6^. Following this report, a separate GWAS replicated the signals at *APOE* and *GBA* (but interestingly, not *SNCA*) in 828 cases and 82,035 controls ^7^. More recently, a GWAS was published merging PDD and DLB cases, with a total of 2,981 cases and 4,391 controls showing associations at *GBA, SNCA*, and *APOE* (presumably driven by DLB cases) as well as two novel signals at *BIN1* and *TMEM175* ^8^. These two additional signals have also previously been associated with AD or PD^9,10^. However, polygenic risk scores (PRS) for AD and PD are not strongly predictive of DLB ^11^, even though these have around 70% accuracy in predicting their respective diseases ^9,10^ and reaching up to 84% accuracy in AD ^12^. These data suggest that, despite the overlaps, DLB has a unique genetic architecture.

DLB has a different incidence in males vs. females, suggesting a sex-specific genetic component ^13,14^. However, the effect of genetics on risk for DLB in a sex-stratified manner has not been determined thus far. In AD, some evidence suggests a role for different genetic risk loci in females vs. males ^15,16^, while in PD, a sex-stratified GWAS reported no differences between sexes but showed some sex-specific associations ^17^.

In this study, we meta-analyzed four DLB GWAS cohorts and one cohort of LBD, including a total of 5,119 cases and 20,988 controls, making it the largest cohort assembled for any of the Lewy body dementias. We dissected the demographic stratification and potential functional implications of these results. We reviewed the polygenicity of the trait and compared it to AD and PD, as well as other common polygenic traits.

## Methods

### Study cohorts

Cohort 1 consisted of the samples previously reported in ^6^ (1,191 DLB cases and 3,723 controls), with the exclusion of 25 samples due to updated diagnoses or cryptic relatedness, which were re-calculated together with samples from Cohort 2.

Cohort 2 comprised 1,166 DLB cases and 5,831 controls and was genotyped on Illumina’s HumanOmniExpress chip (Supplementary Table 1). Control samples were obtained from the Cutaneous Melanoma GWAS Combining Multiple Populations and Risk Phenotypes (phs001868.v1.p1) and the National Cancer Institute (NCI) Head and Neck Cancer Study (phs001173.v1.p1). QC was conducted in the same manner described for Cohort 1, removing variants with a call rate below 95% and GenTrain scores below 0.7.

Both Cohorts 1 and 2 were lifted over to hg38 and prepared for imputation using the tool from the McCarthy group available on (https://www.well.ox.ac.uk/~wrayner/tools/). Imputation was performed on the TOPMed platform ^18^. Samples of non-European ancestry were excluded via PCA analysis with 1000 Genomes, and individuals with relatedness above a 0.04 KING cut-off were excluded. SNPs deviating from Hardy-Weinberg equilibrium (5e-8), with a minor allele frequency below 0.001, and genotyped in less than 99% of samples were also excluded. The majority of cases in Cohorts 1 and 2 were pathologically diagnosed using the 2005 McKeith criteria ^19^, including individuals with an “intermediate” or “high” likelihood of meeting diagnostic criteria for DLB (n = 1,503).

Cohort 3 summary statistics comprised 355 clinical DLB cases and 6,318 controls, before quality controls, from the Spanish GR@ACE/DEGESCO Study ^20^. Samples were genotyped using the Axiom 815K Spanish biobank array and imputed on the TopMed platform. Before imputation, SNPs with missingness above 0.025 and a Hardy-Weinberg equilibrium p-value of less than 1e-15 were excluded. Samples with missingness greater than 0.02, excessive heterozygosity rates (F_HET > interval mean +/- 3sd), and outside +/- 3 median absolute deviation of the 1000 Genomes European population were also excluded to control for population outliers and genotyping errors. Samples with discordant sex assignments were also excluded. After excluding these samples, any SNPs genotyped in less than 95% of samples were also removed.

Cohort 4 summary statistics comprised 594 DLB cases and 2,112 controls, before quality controls, from the European E-DLB consortium. A subset of these samples was described previously ^7^. These samples were genotyped on the Illumina Infinium Omni Express-24 v1.1 platform. QC parameters for this cohort followed those described in the original publication except imputation, which has been updated to use the TopMed platform.

All four cohorts were harmonized using the sumstat-harmoniser from Open Targets (https://github.com/opentargets/genetics-sumstat-harmoniser) to correct for allele discrepancies between datasets. This tool was run using the TopMed reference VCF. Post harmonization, the summary statistics were also annotated using the dbSNP reference VCF for GRCh38.

The replication cohort was obtained from a recently published association in LBD ^8^. It was QCed together with cohorts 1 and 2 to ensure no sample overlap. PLINK IBD analysis of relatedness revealed 784 samples with pi-hat values above 0.8 indicating sample overlap. These samples were excluded from the replication cohort leaving 1,826 cases and 1,920 controls (Table 1). The cases in the replication cohort comprise diagnoses of DLB and PDD, while the discovery cohorts focus on DLB exclusively.

**Table 1:** Discovery and replication cohorts

|  |  | **Total** | **Males** | **Females** | **Neuropathologically diagnosed** |
| --- | --- | --- | --- | --- | --- |
| **Cohort 1** | cases | 1191 | 710 | 481 | 936 |
|  | controls | 3723 | 1745 | 1978 | . |
| **Cohort 2** | cases | 1166 | 711 | 455 | 570 |
|  | controls | 5831 | 3192 | 2639 | . |
| **Cohort 3** | cases | 355 | 123 | 232 | 0 |
|  | controls | 6603 | 3382 | 3221 | . |
| **Cohort 4** | cases | 581 | 356 | 225 | 0 |
|  | controls | 2911 | 1490 | 1421 | . |
| **Meta-Analysis** | cases | 3293 | 1900 | 1393 | 1506 |
|  | controls | 19068 | 9809 | 9259 | . |
| **Replication** | cases | 1826 | 1131 | 674 | 1065 |
|  | controls | 1920 | 972 | 948 | . |
| **Meta-Analysis + Replication** | cases | 5119 | 3031 | 2067 | 2571 |
|  | controls | 20988 | 10781 | 10207 | . |

### Meta-analysis

Analyses of the individual datasets were conducted using logistic regression implemented in PLINK v1.9, using covariates for sex and population stratification. Meta-analysis was also conducted in PLINK using the log-scale model, adjusting for unequal case-control cohorts (effective sample size = 4/(1/cases + 1/controls)). This association was also performed, including only males and females separately for the sex-stratified results. The Bonferroni threshold for genome-wide significance was 5.0 × 10^−8^. A conditional analysis was performed for each GWAS locus significantly associated with disease by the addition of each respective index variant to the covariates. Quantile–quantile plots revealed minimal residual population substructure, as estimated by the sample-size-adjusted, genome-wide inflation factor λ_1,000_ of 1.01 (Supplementary Fig. 1). Power calculations were performed using the genpwr package in R using a logistic model.

### Heritability analysis

We estimated the proportion of heritable SNPs in the meta-analysis results using LD score regression ^21^. Local heritability estimates were calculated using LAVA ^22^. LD blocks for the local heritability were obtained using the predefined regions provided by LAVA. For PD and AD, we used publicly available summary statistics ^9,10^.

### Genetic correlation

The genetic correlation analysis was performed using publicly available summary statistics for PD ^10^ and AD ^9^, which include the UK biobank dataset. Regression coefficients were calculated using LD score regression (LDSC) ^21^. We used the IEU Open GWAS database for other traits, which contains over 10,000 curated, quality controlled, and harmonized complete GWAS summary datasets ^23^. Traits were filtered to only look at IEU-A datasets (datasets curated for the MR-Base tool) ^24^. Only summary statistics including more than 2,000 samples with data from males and females of European ancestry were included. Datasets were then munged and regressed using LDSC.

### Gene-set analyses

Genome-wide gene-based association (burden) and pathway analyses were performed using MAGMA ^25^. For burden association, variants from the meta-analysis results were assigned to genes based on the overlap of their location with a gene window of 35 kb upstream and 10 kb downstream to include regulatory elements. Variants with MAF < 0.05, present in at least 3 out of the four cohorts and annotated with either missense or predicted high impact, were included in a model to assess gene burden. The association between variants in gene units and DLB was tested by mean-SNP association. The MAGMA p-values were corrected for multiple testing by adjusting for the total number of genes. For gene-set association, we included all variants with MAF greater than 0.01 that were present in at least three discovery datasets. Pathways were derived from MSigDB available gene sets, including Gene Ontology, KEGG, REACTOME, BIOCARTA, and the hallmark pathways (http://www.gsea-msigdb.org/gsea/msigdb/index.jsp). We used Bonferroni correction to determine enrichment.

### Phenome-wide association scan

We used PHESANT—PHEnome Scan ANalysis Tool ^26^ to perform an automated phenome scan in the UK Biobank (UKBB). We derived polygenic risk scores from the meta-analysis results using PRSice-2 ^27^ and applied those to the entire UKBB genotyping dataset (filtering for UKBB-described European ancestry). We defined quantiles from the PRS distribution with individuals in the 5% lowest PRS and the 5% highest PRS, as described in ^28^. Phenotypes with more than 20% missing answers were filtered out. We adjusted for sex, age at recruitment, Townsend deprivation index at recruitment, genotype measurement batch, and the first ten principal components. In addition, we considered phenotype categories with a minimum size of 200 answers and converted fields with multiple instances to categorical (multiple) fields as implemented in PHESANT. The same procedure was repeated for sex-stratified meta-analyses and applied to males and females separately.

## Results

### Meta-analysis

The four discovery cohorts were meta-analyzed using a fixed-effects model implemented in plink2.0. The model was selected given the strict inclusion-exclusion criteria set for each cohort, such as filtering out those with strong evidence of originating from a non-European population, diagnostic criteria, adjustment for age, plus similar sex distributions in the full analysis and later, sex stratification. Because of this, we expect each cohort to be estimating the same common effect size. Despite more than doubling the sample size from previous analyses, no genome-wide associations were detected beyond those previously reported in *GBA, SNCA* and *APOE* (Fig. 1). As expected, all three loci replicated in the replication cohort. The *STX1B/BCL7C* locus on chromosome 16 showed suggestive significance, and when performing a joint analysis including discovery and replication cohorts, it showed a genome-wide significant p-value (Table 2).

**Figure 1:**
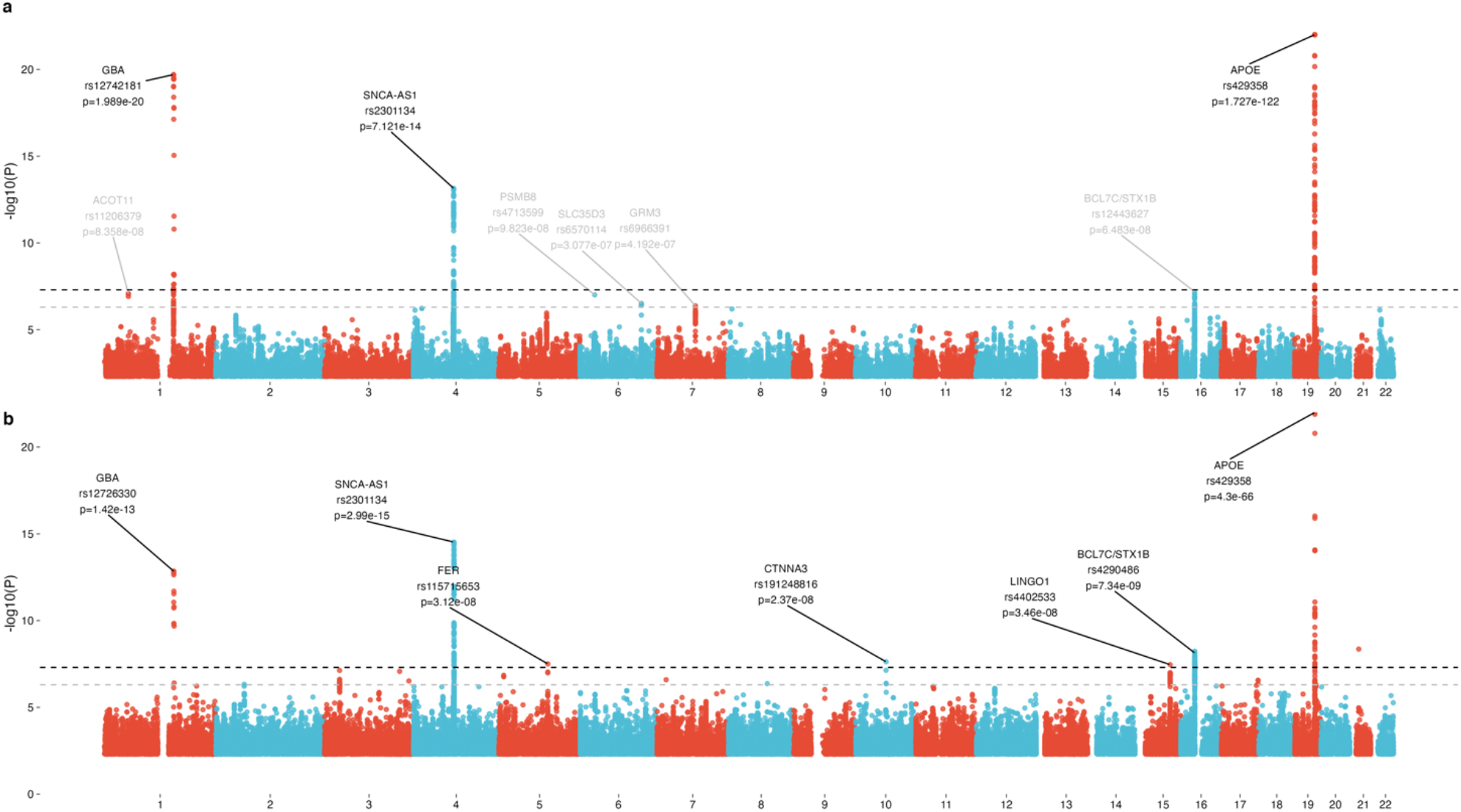
Manhattan plots of the full DLB meta-analysis (a) and the meta-analysis including only pathologically diagnosed cases (b). The dashed black line indicates the genome-wide significant threshold at P < 5×10-08, while the dashed grey line indicates suggestive threshold of 5×10-07. Note that the y-axis has been capped at P=1×10-22 for better visualization of signals below APOE. Please refer to labels for maximum p-values of APOE.

**Table 2:** Top SNPs from meta-analysis.

| Chr | Pos | SNP | Locus | A1 | AX | P | Beta | A1_Freq | Replication_P | Joint_P | Joint_BETA |
| --- | --- | --- | --- | --- | --- | --- | --- | --- | --- | --- | --- |
| 1 | 155143675 | rs12742181 | GBA | T | C | 1.99E-20 | 1.0838 | 0.011067 | 0.000616 | 1.99E-20 | 1.0838 |
| 4 | 89837794 | rs2301134 | SNCA-AS1 | G | A | 7.12E-14 | 0.2311 | 0.507666 | 0.000407 | 1.84E-16 | 0.2171 |
| 16 | 30985551 | rs12443627 | BCL7C/STX1B | C | G | 6.48E-08 | 0.1726 | 0.587742 | 0.078258 | 3.20E-08 | 0.1507 |
| 19 | 44908684 | rs429358 | APOE | C | T | 1.73E-122 | 0.8976 | 0.142765 | 1.62E-30 | 1.90E-150 | 0.863 |

Results show p-values and effect sizes from fixed effects model. Joint\_P and Joint\_BETA refer to the joint analysis of discovery and replication.

Given the mentioned diagnostic challenges with DLB and the substantial overlap we see between genetic signals with AD and PD, we also assessed the association using only individuals with both clinical and pathological diagnoses (Fig 1b). This analysis yielded the previously detected peaks at *GBA, SNCA, APOE*, and *STX1B/BCL7C* and additional genome-wide significant loci near *FER, CTNNA3, LINGO1*.

Recently, two loci have been suggested to be associated with LBD - *TMEM175* and *BIN1* ^8^. Using the reported effect size and allele frequency for each of the leading variants at these loci, despite being adequately powered to detect them (0.988 and 0.993 power, respectively), we did not identify significant associations in either of these loci. We also did not detect common variants at the *LRP10* or *PLCG2* loci, which were also reported to modulate risk for DLB recently ^29,30^. The reported variant in *PLCG2* is of lower frequency, so it was excluded in our minor allele frequency filtering. However, frequencies in cases and controls in each tested cohort show no evidence of a difference, as indicated by the overall p-value calculated using the same methods above, without a frequency filter (rs72824905; p-value = 0.1566; BETA = -0.2821).

The conditional analysis did not yield any additional genome-wide significant peaks. However, there was some evidence for independent signals at the *GBA* and *SNCA* loci (Supplemental Fig. 2).

### Sex-stratified meta-analysis

Cohorts were stratified by sex and meta-analyzed as described in the discovery meta-analysis section. This yielded a male cohort of 1,900 cases and 9,809 controls and a female cohort of 1,393 cases and 9,259 controls. Significant associations at *GBA* and *SNCA* were only present in males (Fig. 2). Stratifying these results by sex reduces some of the heterogeneity in the overall meta-analysis. However, there is still high heterogeneity within the most significant SNPs in males, suggesting perhaps population or diagnostic heterogeneity (Supplementary Fig. 3).

**Figure 2:**
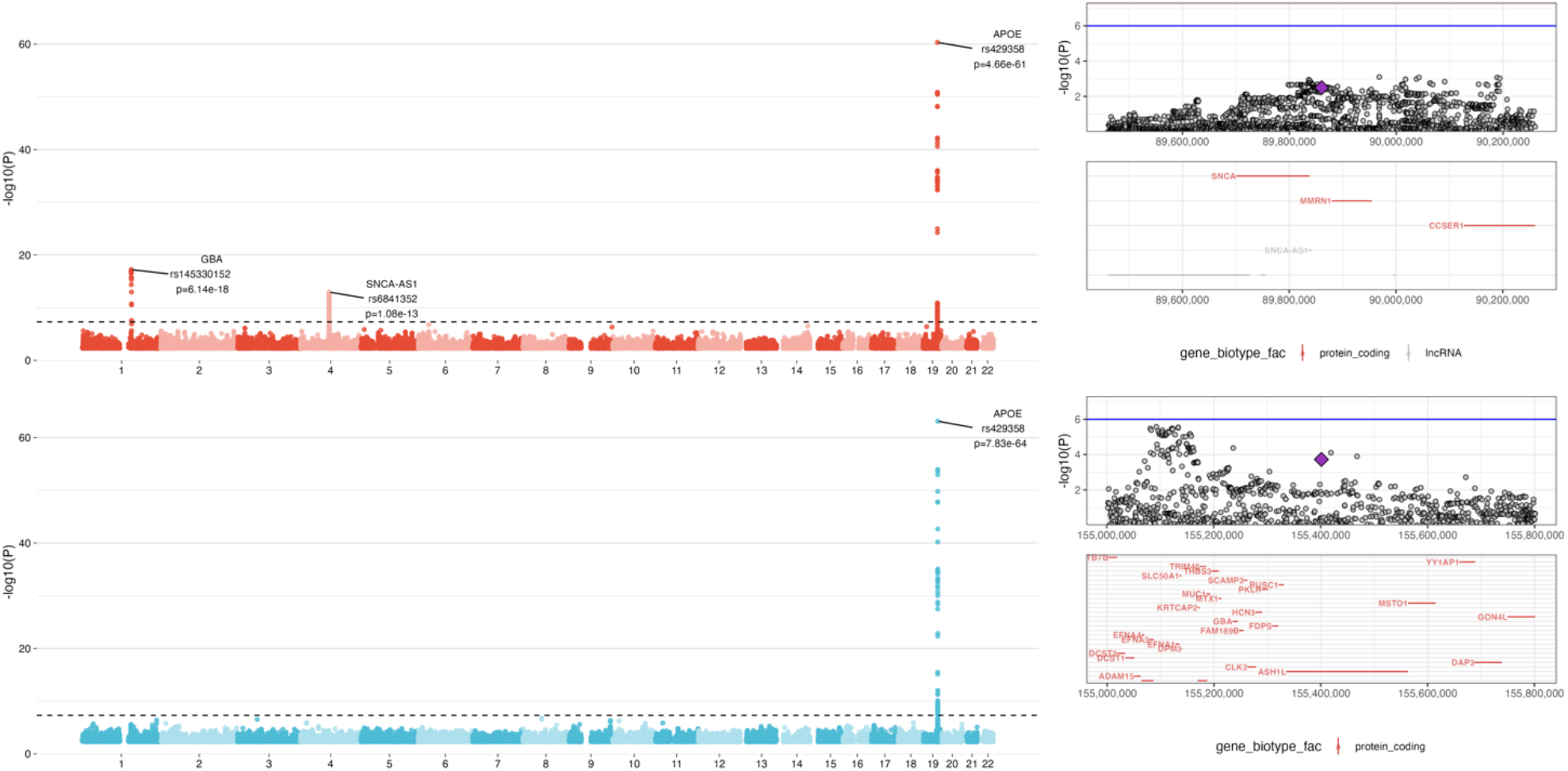
Meta-analysis stratified by males (a) and females (c) and locus zoom plots of GBA (b) and SNCA (d) loci in females. The purple diamonds in locus zoom plots indicates the top associated SNPs from the males GWAS.

In addition to not showing significance in females, the *GBA* and *SNCA* loci have significantly different effect sizes between sexes with no overlap in their confidence intervals (Table 3).

**Table 3:** Significant loci in the sex-stratified analysis

| Chr | Pos | SNP | Locus | A1 | A2 | Male_BETA | Male_SE | Male_P | Female_BETA | Female_SE | Female_P |
| --- | --- | --- | --- | --- | --- | --- | --- | --- | --- | --- | --- |
| 1 | 155401328 | rs145330152 | GBA | C | A | 1.1848 | 0.1373 | 6.14E-18 | 0.6514 | 0.1745 | 0.0001891 |
| 4 | 89860504 | rs6841352 | SNCA-AS1 | T | G | 0.3098 | 0.0417 | 1.08E-13 | 0.1362 | 0.0462 | 0.003199 |
| 19 | 44908684 | rs429358 | APOE | C | T | 0.8512 | 0.0516 | 4.66E-61 | 0.9648 | 0.0572 | 7.83E-64 |

For rs145330152 and rs6841352, there is significant evidence that these effects differ by sex (p = 0.016 and p = 0.005, respectively). For rs145330152, males had an effect ∼1.8 times larger than females (95% CI:1.15 - 2.5-fold increase) and 2.27 times larger for rs6841352 (95% CI:1.38 - 3.17-fold increase). There was no difference between the sex-stratified datasets at the *APOE* locus (rs429358; p=0.14, 95% CI 0.95 - 1.31). P-values were calculated using a z-test as described in ^31^.

### Low Frequency Variants

A gene burden analysis was conducted to assess the role of low frequency variation (MAF < 0.05) (Fig. 3). Only *GBA* was significantly associated with DLB, with three variants leading the signal (NM_000157.4:p.(Glu365Lys), p.(Asn409Ser) and p.(Thr408Met)).

**Figure 3:**
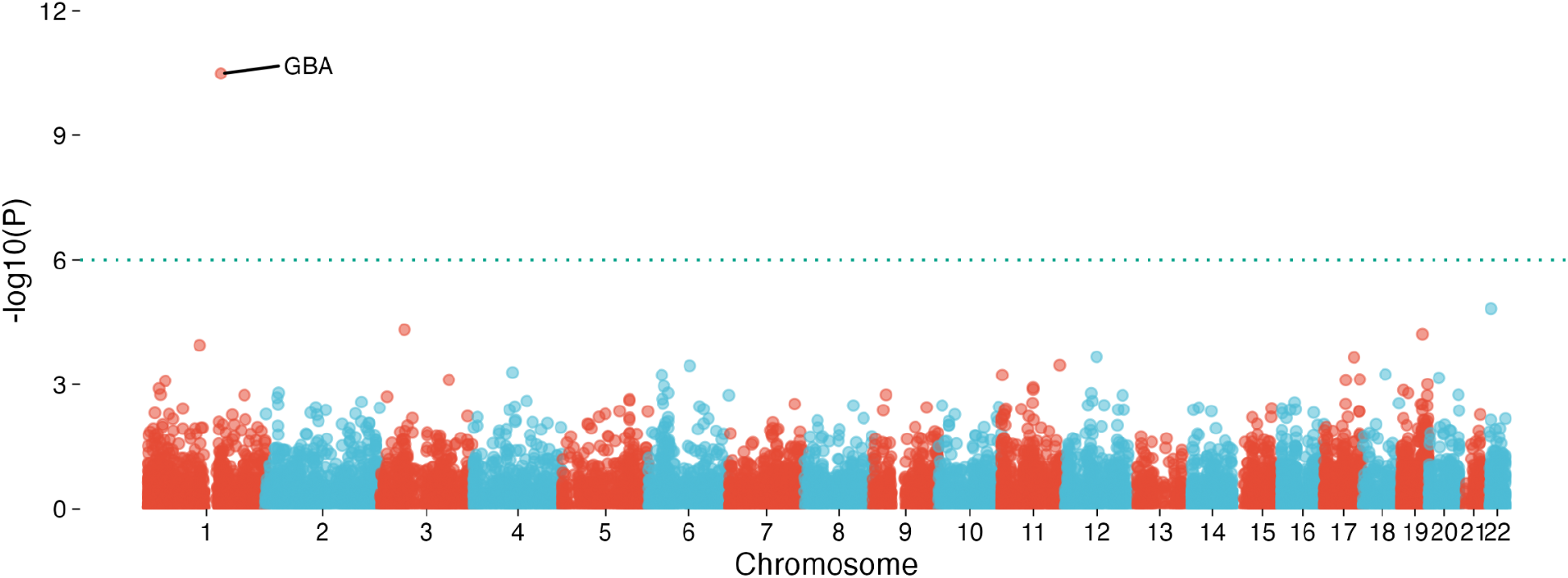
Genome-wide plot of gene burden results. Shown here are associations from variants with Ensembl VEP annotation of either missense or high impact, and those variants with MAF below 0.05 and found in at least ¾ of the meta-analysis cohorts

### Heritability

Using the full meta-analysis data, we estimated the heritability of DLB on the liability scale using a disease prevalence of 0.004 ^32^ to be 13.11% (SE = 0.03). To obtain a fine-mapping of the overall heritability in DLB, we applied the tool LAVA, using predefined blocks of LD to identify regions of high heritability in the genome. Unsurprisingly, *APOE* and *GBA* were in the two highest heritability regions, with highly significant heritability estimates (p= 6.45E-123 and p= 7.13E-21, respectively). Interestingly, the third highest region was on chromosome 13, encompassing three genes not previously associated with DLB: *EFNB2, ARGLU1* and *FAM155A* (Table 4). This result is in line with our previous analysis showing a disproportionate high heritability on chromosome 13 despite identifying no genome-wide significance results ^6^, using an orthogonal method (Fig. 4a).

**Table 4:**
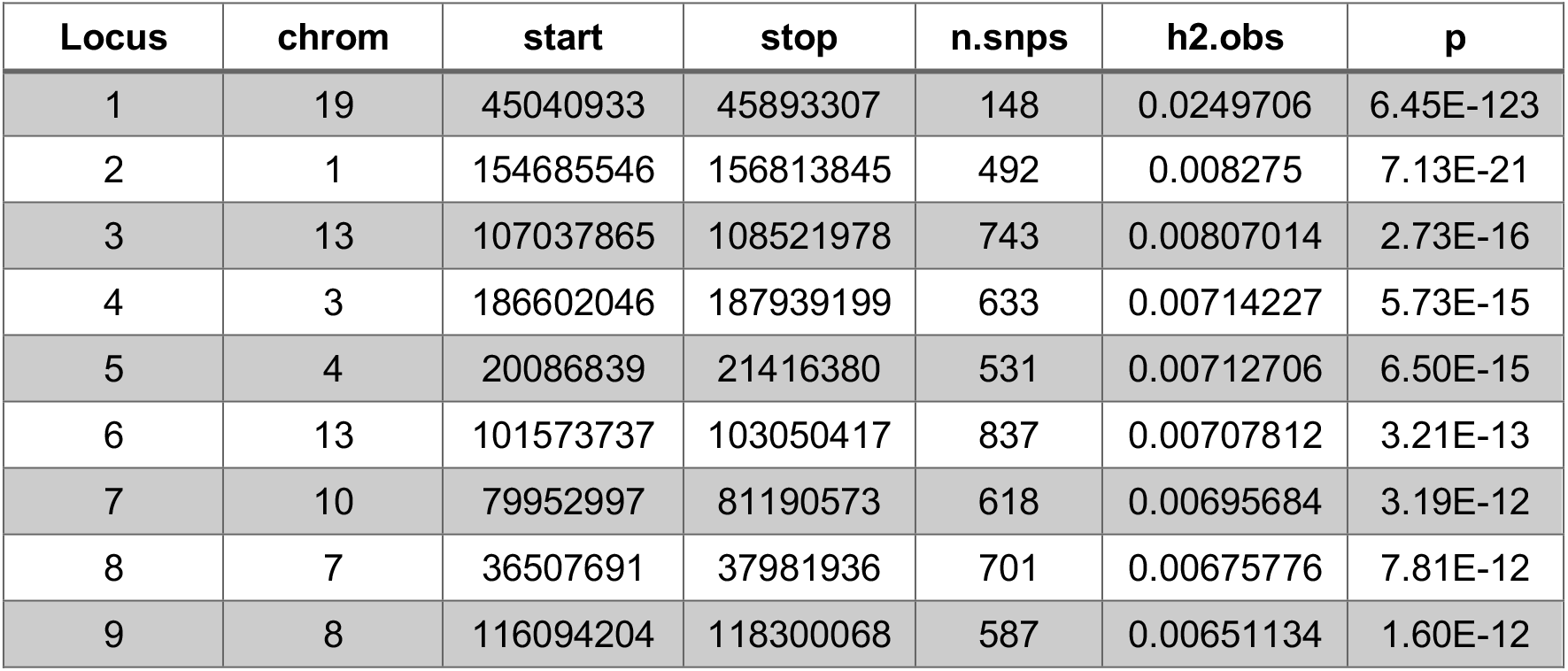
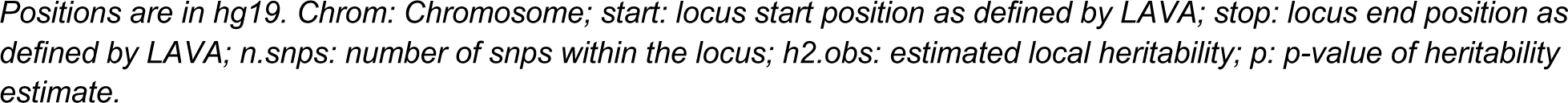
LAVA loci with the highest DLB heritability

**Figure 4:**
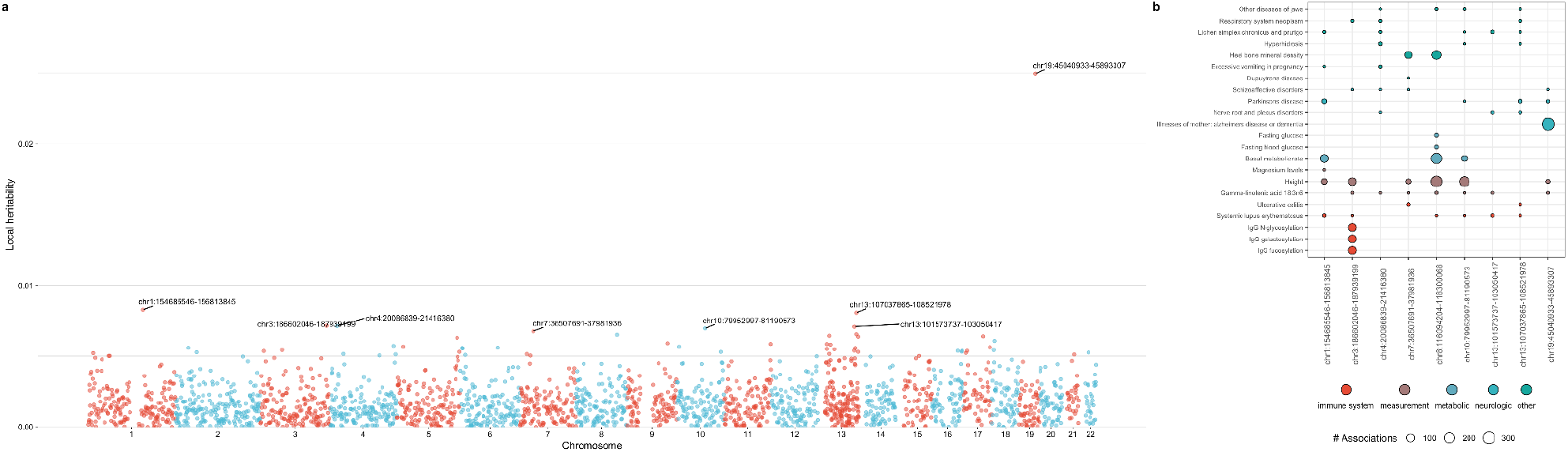
Genome-wide plot of LAVA h2 values in DLB meta-analysis (a). Positions are in hg19. Regions with the largest number of associations as detected by PhenoScanner within these top heritable loci (b). Trait groupings based on OLS ontology search.

To identify potential pleiotropic associations between the high heritability regions and other complex traits, we interrogated the PhenoScanner database ^33^. Associations within these regions include IgG modifications in the chr3:186602046-187939199 locus, as well as associations relating to metabolism and growth in the chromosome 8 locus. Notably, the *SNCA* locus is not among the regions with the highest heritability (Fig. 4b).

### Genetic Correlation

We determined the genetic correlations between DLB and traits reported in the MRCIEU GWAS catalog using LDSC. As previously reported, we saw a positive correlation between DLB and “Years of schooling”. This correlation seems to be driven exclusively by the male genetic results. Most negatively correlated traits included multiple sclerosis, phenotypes relating to smoking use, type 2 diabetes, and anthropometric traits corresponding to a greater body mass (Fig. 5). When performing the genetic correlation estimate between males and females, we identified a non-significant correlation (rg=0.30; se=0.28; p=0.28), further supporting a sex-specific genetic architecture.

**Figure 5:**
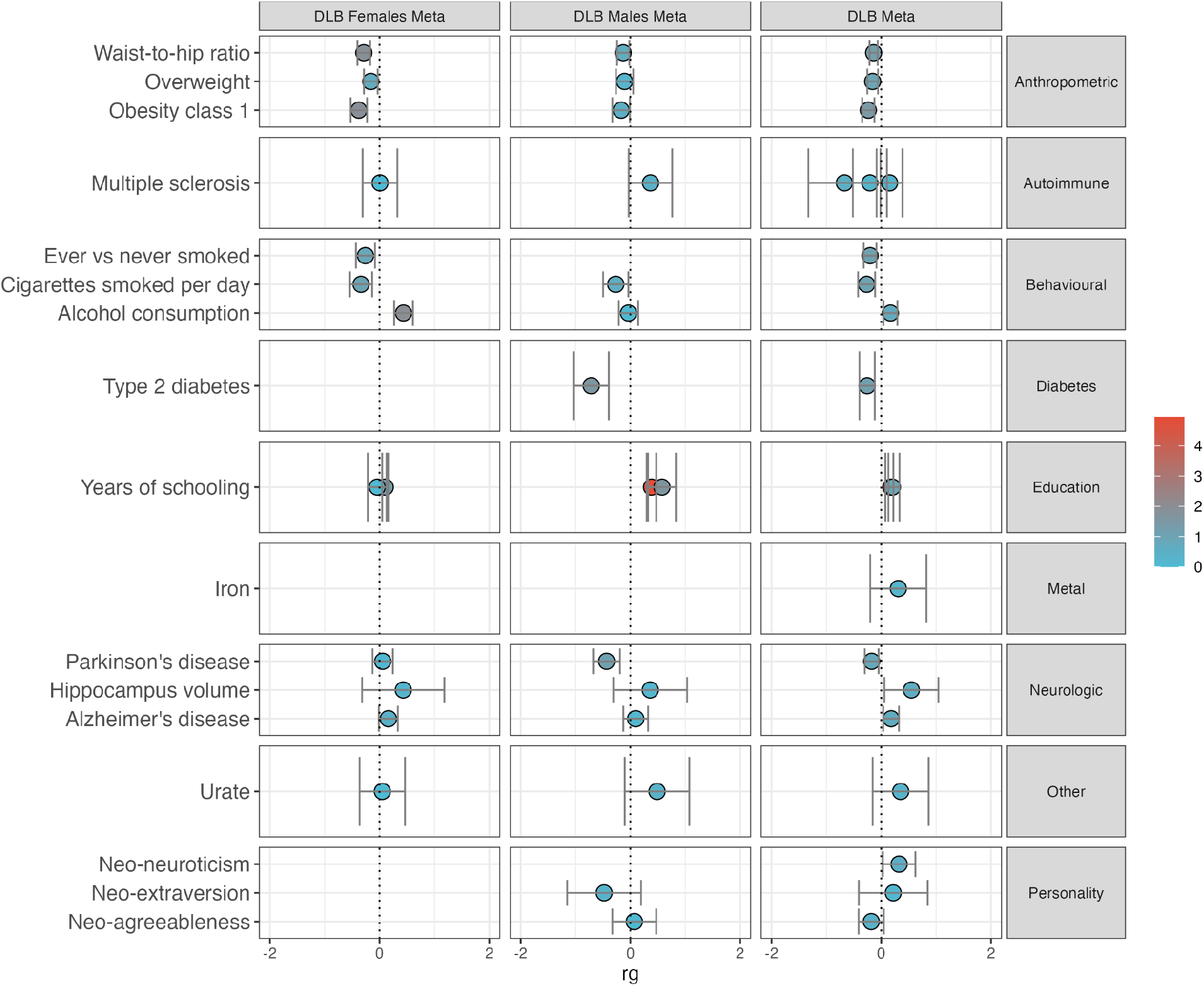
Top genetic correlations between MRCIEU traits. Color indicates -log10(pvalue).

To dissect the specific loci that underpin genetic correlations between common neurodegenerative diseases, we mapped each of the previously identified GWAS hits in AD, PD and DLB to the blocks predefined by LAVA for the European population mentioned above. We then estimated the level of genetic correlation at each locus between the diseases. Interestingly, some loci, such as the HLA region, *TMEM175, ECHDC3*, and *ADAMTS4* showed strong correlation between AD and PD.

Additionally, several AD and PD loci showed strong correlations with DLB, with *GBA, APOE, BIN1*, and *SCIMP* displaying the most significant p-values (Fig. 6).

**Figure 6:**
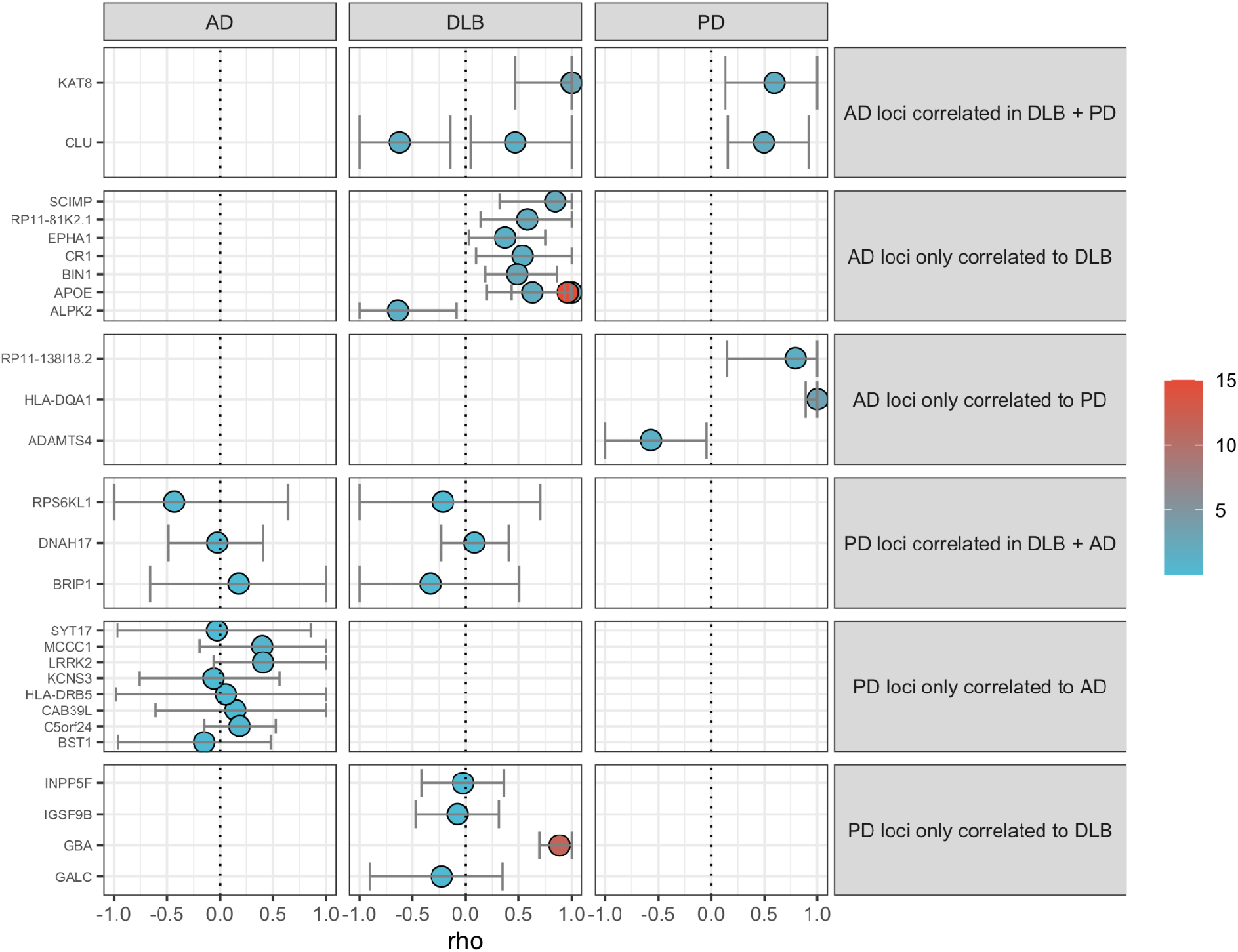
AD and PD GWAS loci correlations across the three most common neurodegenerative diseases. Note that genes with multiple correlations reflects multiple LAVA loci being mapped to the same GWAS association locus. Color indicates -log10(p-value). APOE p-value has been capped at 1e-15 to capture variation at less significant loci. The APOE region has a p-value of 5.45E-132 between AD and DLB.

### Gene-set Enrichment

Gene-set enrichment revealed the strongest enrichments in aspartic-type endopeptidase activity, *APP* catabolic processing, and cell surface-related proteins (Fig. 7). These two most significant associations were mainly driven by the association at chromosome 1, upstream of *GBA*.

**Figure 7:**
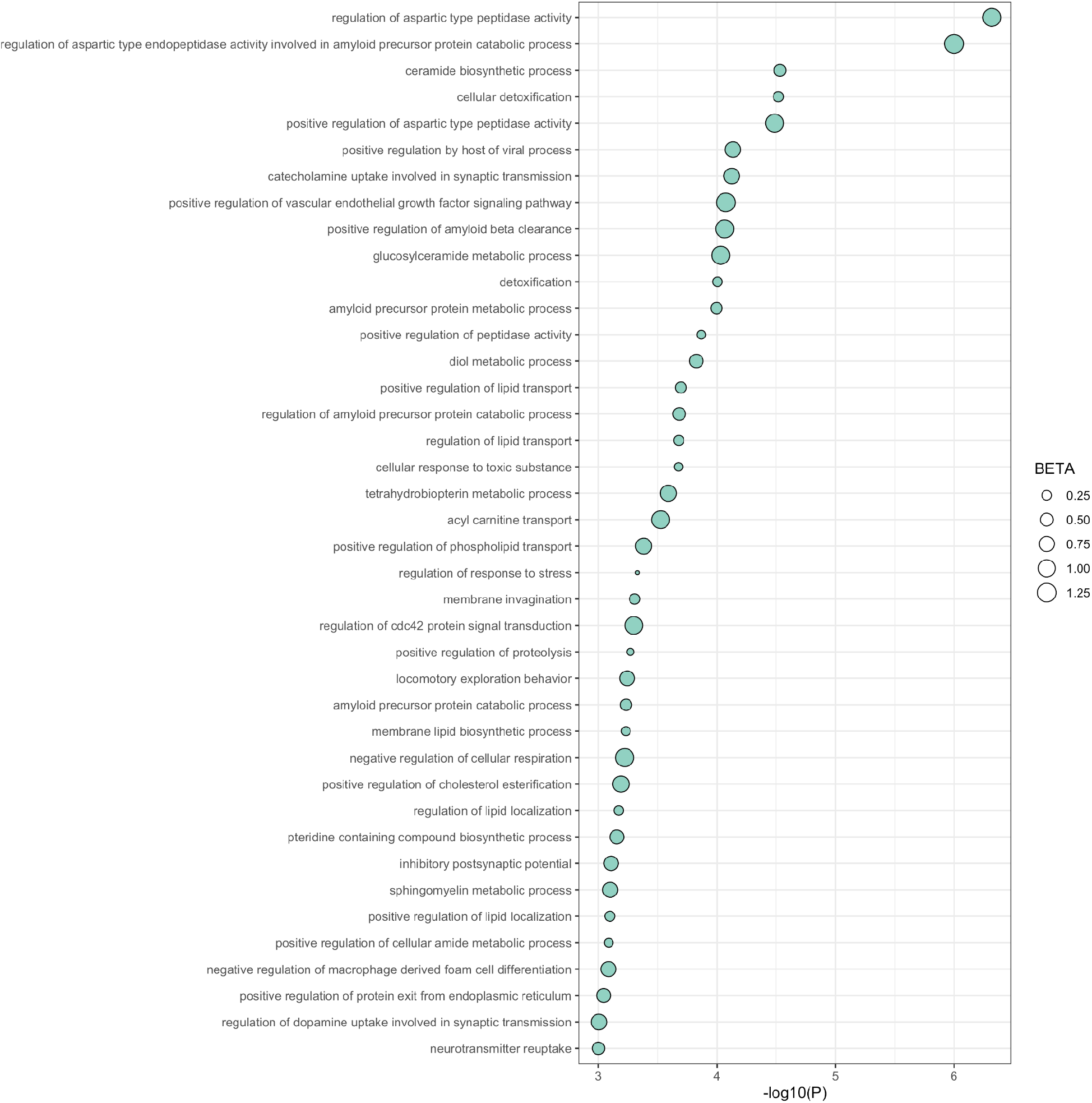
Summary of top (P<0.001) associated GO biological processes genesets in the meta-analysis. The size of points reflects beta values.

### Phenome-wide association study

We investigated the relationships between weighted PRSs constructed from the meta-analysis results and phenotypes or traits available from the UK Biobank. This analysis showed significant associations with family history of dementia, apolipoprotein, and cholesterol levels (Fig. 8). Many of these associations can be directly attributed to the strong effect *APOE* has on various measured phenotypes, while others, such as anthropometric measurements, are unlikely to be directly driven by *APOE* genotype.

**Figure 8:**
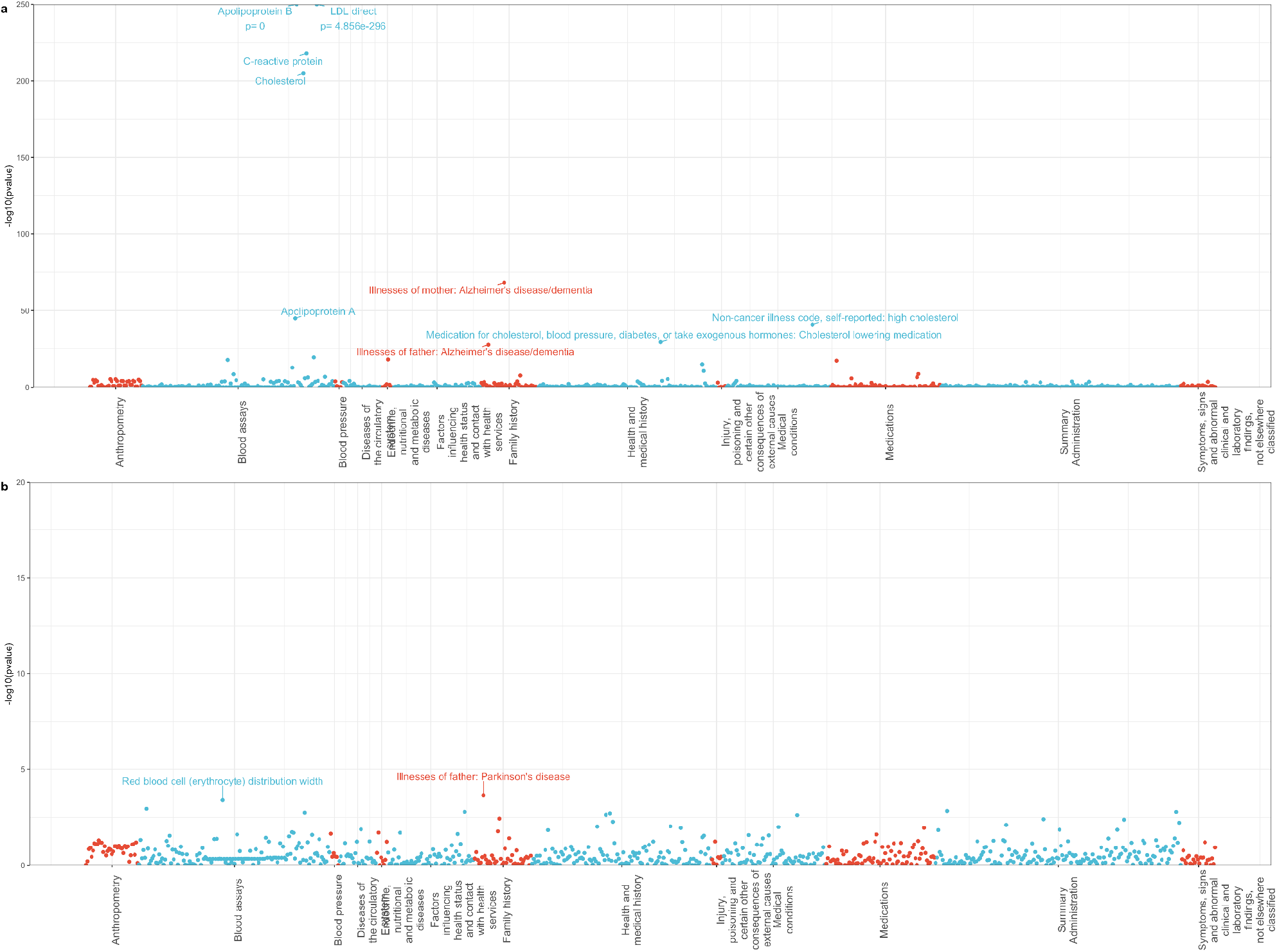
PheWAS summary generated via PHESANT software in the DLB meta-analysis. Results shown stratified by PRS extremes generated with (A) and without (B) the APOE region to better understand this strong effect on risk.

Phenotypic characterization of the male and female PRSs was limited due to low sample size in these cohorts and yielded no significant associations. However, the impact of these different genetic profiles is particularly appreciable in the effect sizes for AD and PD-related risk. Males with high male-DLB genetic risk have an increased incidence of familial PD, but only paternally (Fig. 9). Likewise, females with high female-DLB genetic risk have an increased incidence of maternal AD, while this trait was not among the most significant in males (Supplementary Table 2).

**Figure 9:**
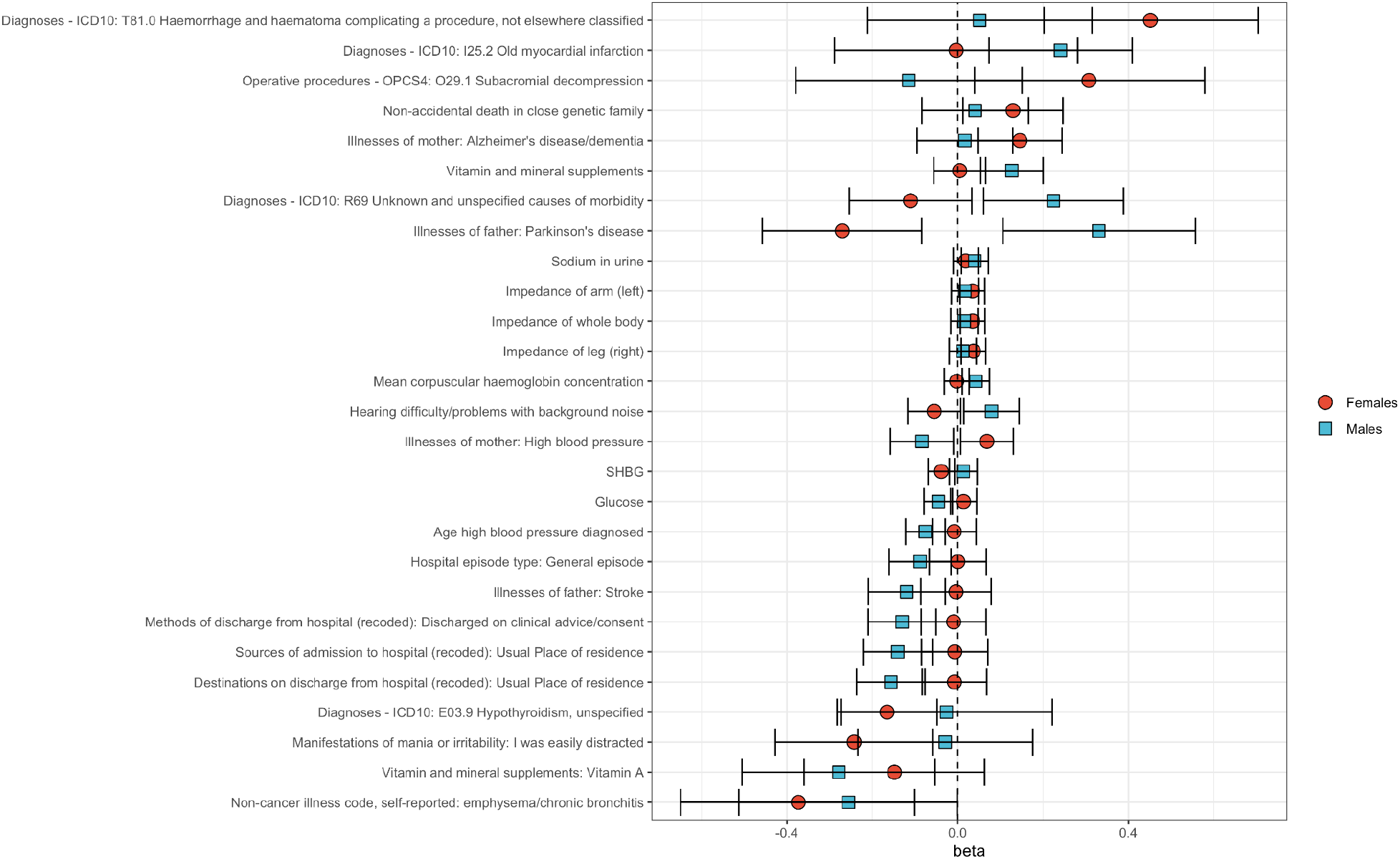
Male vs female differences in PheWas results. Since no phenotypes were genome-wide significant, only betas were plotted to visualize areas of greatest differences between males and females based on the respective sex-stratified GWAS previously described.

## Discussion

This is the largest genome-wide study in DLB, using a cohort that doubles the size of previous studies. Despite the substantial increase in cohort size, we do not observe additional associations to those previously identified at *GBA, SNCA*, and *APOE*. The genetic architecture of DLB may be comprised of associations with lower frequency variants that we are still not powered to identify with the current cohort size, as previously suggested ^11^. It is also possible that the clinical heterogeneity and difficulty in accurately diagnosing DLB contribute to these findings. As an example of this heterogeneity, we identify a sex difference in the genetic results suggesting that genetics are a more prominent risk factor for DLB in males than females. Using cohorts of comparable size, we failed to identify associations at *GBA* and *SNCA* in females. This finding may also partially contribute to the higher prevalence of DLB in males compared to females and is an important aspect to consider when selecting cohorts for clinical trials. A recent report on AD has shown that using sex-dependent autosomal effects improves the predictive ability of the genetic results ^34^. Additionally, it has been recently shown that there are sex-specific associations with cerebrospinal fluid biomarkers in DLB ^35^. One of the results was lower levels of CSF alpha-syn in females, a finding that would be in agreement with a lack of association at this locus, assuming the mechanism of association is through an increase in expression of *SNCA*, as previously demonstrated ^6^. Lastly, sex is an important driver of brain pathology and clinical phenotypes of DLB, as recently stated in both clinical and neuropathologically-confirmed samples ^36^. Together, these findings add support to a disease pathobiology that has some sex-differences.

The absence of an association at *GBA* in females is interesting as this locus has shown heterogeneous results in different publications. A study in an Italian cohort of DLB observed 79% of *GBA* mutation carriers were male. Notably, this sex stratification was less pronounced in PDD (64%) and absent in PD (49%) in the same study ^37^. On the other hand, Blauwendraat and colleagues reported that *GBA* has a more significant p-value in females compared to males in their sex-stratified GWAS in PD ^17^. Lastly, in a Spanish GWAS in PD, *GBA* showed no association with disease ^38^, further highlighting the heterogeneity present at this locus in PD.

One approach to minimize heterogeneity is using cases with the highest diagnostic confidence. For DLB, that means including individuals with both clinical and pathological diagnoses of the disease. Performing a GWAS using this subset of cases highlights the same three loci as associated with disease in the larger cohort and shows four other loci surpassing Bonferroni. The chromosome 16 locus (*BCL7C*/*STX1B*) had been previously highlighted as associated with DLB and is also associated with both PD and AD. The *LINGO1* locus has been previously associated with essential tremor ^39^, and given that there is a recognized increase in dementia among essential tremor patients ^40^, it is plausible these results reflect the same association reported here. *CTNNA3* encodes alpha-T-catenin and is a binding partner of β-catenin, which interacts with *PSEN1* ^41^. It was also shown to be associated with the level of plasma Aβ42 in LOAD ^42^, suggesting a role in dementia. The *FER* locus has not been associated with neurodegenerative phenotypes, although associations have been reported with anthropometric traits ^43^. The loci identified in the pathological subset analysis should be interpreted with caution. Although they are genome-wide significant, they are borderline significant, and we are unable to properly replicate them in independent datasets since these are not available.

An important locus in PD is the one encompassing *LRRK2*; mutations in this gene are one the most common causes of PD and common variants are associated with sporadic forms of disease. This makes *LRRK2* an interesting therapeutic target in PD. Interestingly, we do not see a signal at this locus (lowest p-value at *LRRK2* was rs138538499; p=2.76E-3), with the nearby by locus *CNTN1*, which we previously reported contained a suggestive signal, showing a p-value of p=8.29E-6. These data suggest that therapeutic approaches targeting *LRRK2* might be less effective in DLB than PD.

The increase in sample size also allows us to attempt to replicate recently published associations. For example, a recent report ^30^ detailed how a low-frequency variant in *PLCG2* is associated with DLB. We did not see that association in our cohort that uses about 4x the number of cases.

Similarly, two loci were recently reported to be associated with LBD (*TMEM175* and *BIN1*) ^8^. Although the inclusion criteria for LBD are different than that for DLB (the former includes PD with dementia, the latter does not), we do not see evidence of association for either locus in our data. It is plausible that these are associations driven by PDD cases, reinforcing the importance of properly characterizing cases and datasets, particularly when studying diseases with clinical and diagnostic heterogeneity.

To quantify how much of the genetic liability the large meta-analysis can explain, we generated updated heritability estimates. Using our discovery cohort, we estimate the liability-scale heritability of DLB as 0.1313 (S.E. 0.0317), which is only slightly lower than the heritability estimated for PD (0.18-0.26) ^10^. We used the recently developed method LAVA in our discovery dataset to narrow down the genome regions harboring high heritability. As expected, the two loci with highest heritability overlap *APOE* and *GBA*. Interestingly, chromosome 13 contains the next highest heritability regions, following previous data suggesting a disproportionally high heritability given the chromosome size ^6^. Using LAVA, we can fine-map that chromosome-wide heritability to specific loci. Since we do not identify significant associations, variants of lower frequency or exceedingly low effects in these loci likely drive these results, highlighting the need for larger DLB cohorts to be studied.

To identify traits that have been associated with loci within our highest heritability regions, we performed a PhenoScanner analysis using these regions. As expected, we identify Alzheimer’s-related traits in chromosome 19 and Parkinson’s in chromosome 1. We also identify several immune-related traits in chromosomes 3 and 13, suggesting a potential overlap between DLB and these phenotypes.

Given the many parallels between DLB, PD, and AD, we sought to use the known genetic risk loci for each of them to identify local correlations using LAVA. Two AD loci are correlated with both PD and DLB - *BCL7C/STX1B* and *CLU*, while no PD loci are strongly correlated with AD and DLB. The *BCL7C/STX1B* locus, which also encompasses the *KAT8* gene, seems to be the only locus with some evidence of association across all three neurodegenerative diseases. One of the genes at the locus, *KAT8*, directly interacts with KANSL1 of the NSL complex ^44^. *KANSL1* has been suggested to be the causative gene driving the *MAPT* signal and has also been associated with AD ^45,46^. In PD, the locus has been associated via GWAS and differential expression data ^47,48^.

We identified significant genetic correlations with other traits and diseases. “Hippocampus volume” showed a positive correlation with DLB, a finding that we do not see in PD or AD, even though this is one of the most affected brain areas in dementia. “Years of schooling” was also positively correlated with DLB, which seems to be driven by males since no significant correlation was observed for females. Our results also show a positive correlation of DLB with AD and a negative correlation with PD, using the publicly available summary statistics for these diseases, although neither of these correlations reached significance. We identified a negative genetic correlation between DLB and type-2 diabetes. Interestingly, a recent report found a similar result for AD, suggesting that therapeutic strategies for these diseases would need to be disparate ^49^.

The PheWas analysis identified several traits associated with the genetic risk from DLB that are largely driven by the strong effect that APOE has in a variety of traits. When excluding APOE from the risk score, there were no significant traits identified. We were unable to identify significant associations with traits in a male and female specific PheWas, likely due to the lower sample size in these cohorts. However, in females there was a trend for maternal AD risk, while in males the risk was associated with paternal PD.

We acknowledge some limitations in our study. First, our study population is of European descent, and the inclusion of genetically diverse samples is critical to improving our understanding of the genetic bases of disease. We use a cohort for replication for which the diagnostic criteria are not the same as those used for discovery. However, we note that no other cohorts are available that could be used to replicate the discovery findings. We have calculated heritability and genetic correlation estimates for our DLB cohort using LDSC. It is known that this method was designed and calibrated for large-scale sample sizes, which suggests that some of the heterogeneity in these analyses may be a consequence of the relatively small cohort size.

The sex-stratified female cohort is smaller than the male cohort and this could underlie the differences in associations between the two cohorts. However, this cohort is well-powered to detect the associations identified in the male stratified cohort with >0.999 and 0.98 statistical power to identify the variants at *GBA* and *SNCA* at an alpha of 5×10^−08^, respectively.

In summary, despite more than doubling the size of previous GWAS in DLB, we do not identify additional common variants associated with this disease in our main discovery dataset. It is plausible that this is because the genetic architecture of DLB comprises lower frequency and/or effect size variants, for which larger, well-defined cohorts are necessary to identify them. It also may be a reflection of unappreciated heterogeneity in these cohorts as a pathologically diagnosed subset yielded an additional four genome-wide significant signals. However, since this is a smaller subset of the cohort, these signals would require independent replication. For the first time, we identify a sex-specific genetic signature in DLB that may contribute to the distinct disease prevalence in males and females. These results also suggest that some of the heterogeneity we observe in our data is driven by the distinct genetic basis of DLB in males and females. Altogether, these findings represent a substantial advance in our understanding of the genetic architecture of DLB and provide a foundation to develop therapeutic interventions as well as improved diagnostics for DLB and dementia more broadly.

## Data Availability

Summary statistics will be deposited in the GWAS catalog

## Contributors

EG, ARo, IdR, ASh, KW, AB, LR, OA, ARu, DA, RG and JB were responsible for study-level analysis. EG, KW, AB, RG, JB were responsible for additional analysis and data management. ARo, ARu, DA, RG, JB were responsible for design and funding. All authors were responsible for critical review and writing of the manuscript.

## Data sharing

Summary statistics from this work will be deposited in the GWAS catalog.

## Acknowledgments

JB is supported by the National Institutes on Aging (R56 #AG070857) and the National Institutes of Neurological Diseases and Stroke (RF1 #NS124848). RG is supported by the National Institutes on Aging (R01 #AG067426). ER is supported by the Canadian Consortium on Neurodegeneration in Aging. We are grateful to the Banner Sun Health Research Institute Brain and Body Donation Program of Sun City, Arizona for the provision of human biological materials. The Brain and Body Donation Program has been supported by the National Institute of Neurological Disorders and Stroke (U24 NS072026 National Brain and Tissue Resource for Parkinson’s Disease and Related Disorders), the National Institute on Aging (P30 AG19610 Arizona Alzheimer’s Disease Core Center), the Arizona Department of Health Services (contract 211002, Arizona Alzheimer’s Research Center), the Arizona Biomedical Research Commission (contracts 4001, 0011, 05-901 and 1001 to the Arizona Parkinson’s Disease Consortium) and the Michael J. Fox Foundation for Parkinson’s Research. This research has been conducted using data from UK Biobank, a major biomedical database (www.ukbiobank.ac.uk) under application number 11036. The authors acknowledge the contribution of data from the studies National Cancer Institute (NCI) Head and Neck Cancer Study, Cutaneous Melanoma GWAS Combining Multiple Populations and Risk Phenotypes and DEMENTIA-SEQ: Genome sequencing in Lewy Body Dementia, Frontotemporal Dementia, and neurologically healthy controls, accessed through the database of Genotypes and Phenotypes (phs001173.v1.p1, phs001868.v1.p1 and phs001963.v1.p1, respectively). GR@ACE cohort: We would like to thank patients and controls who participated in this project. The Genome Research @ Ace Alzheimer Center Barcelona project (GR@ACE) is supported by Grifols SA, Fundación bancaria ‘La Caixa’, Ace Alzheimer Center Barcelona and CIBERNED. We are indebted to Trinitat Port-Carbó legacy and her family for their support of Ace Alzheimer Center Barcelona research programs. Ace Alzheimer Center Barcelona is one of the participating centers of the Dementia Genetics Spanish Consortium (DEGESCO). A.R. and M.B. receive support from the European Union/EFPIA Innovative Medicines Initiative Joint undertaking ADAPTED and MOPEAD projects (grant numbers 115975 and 115985, respectively). M.B. and A.R. are also supported by national grants PI13/02434, PI16/01861, PI17/01474, PI19/01240 and PI19/01301. Acción Estratégica en Salud is integrated into the Spanish National R + D + I Plan and funded by ISCIII (Instituto de Salud Carlos III)– Subdirección General de Evaluación and the Fondo Europeo de Desarrollo Regional (FEDER– ‘Una manera de hacer Europa’). I.dR. is supported by national grant from the Instituto de Salud Carlos III FI20/00215. Some control samples and data from patients included in this study were provided in part by the National DNA Bank Carlos III (www.bancoadn.org, University of Salamanca, Spain) and Hospital Universitario Virgen de Valme (Sevilla, Spain); they were processed following standard operating procedures with the appropriate approval of the Ethical and Scientific Committee. OAR is supported by NIH (RF1 NS085070; U54-NS100693; U01 NS100620; R01 AG056366; U19 AG071754), DOD (W81XWH-17-1-0249), The Michael J. Fox Foundation, The Little Family Foundation, the Mayo Clinic Dorothy and Harry T. Mangurian Jr. Lewy Body Dementia Program at Mayo Clinic, Ted Turner and family with the Functional Genomics of LBD Program and the Mayo Clinic LBD Center without Walls (U54-NS110435). Mayo Clinic is an American Parkinson Disease Association (APDA) Mayo Clinic Information and Referral Center, an APDA Center for Advanced Research, and a Lewy Body Dementia Association (LBDA) Research Center of Excellence.

