## Supplementary Material for "Identification of a sex-specific genetic signature in dementia with Lewy bodies: a meta-analysis of genome-wide association studies"

#### Tables

Supplementary Table 1: Cohort 2 demographics

| Country | N cases | N Pathologically diagnosed | Male:Female Ratio | mean AAO | mean AAD |
|---|---|---|---|---|---|
| **France** | 83 | 0 | 0.729 | 70.012 | NA |
| **Italy** | 76 | 0 | 1.171 | 74.633 | NA |
| **Spain** | 71 | 39 | 1.290 | 69.284 | 76.974 |
| **Sweden** | 17 | 0 | 3.25 | NA | NA |
| **UK** | 170 | 67 | 2.542 | 74.263 | 84.935 |
| **USA** | 749 | 464 | 1.592 | 65.654 | 78.513 |

Supplementary Table 2: Most significant traits in the sex-specific PRS extreme analysis using PHESANT PheWAS.

| Trait | Dataset | Females_beta | Females_pvalue | Males_beta | Males_pvalue |
|---|---|---|---|---|---|
| Diagnoses - ICD10: T81.0 Haemorrhage and haematoma complicating a procedure, not elsewhere classified | Female | 0.451351213 | 0.000421335 | 0.051427236 | 0.700958368 |
| Illnesses of mother: Alzheimer's disease/dementia | Female | 0.145696683 | 0.003817525 | 0.017015231 | 0.765773158 |
| Diagnoses - ICD10: E03.9 Hypothyroidism, unspecified | Female | -0.165443639 | 0.005744626 | -0.026258722 | 0.835298292 |
| Non-cancer illness code, self-reported: emphysema/chronic bronchitis | Female | -0.373403662 | 0.00756735 | -0.255756069 | 0.049979716 |
| Manifestations of mania or irritability: I was easily distracted | Female | -0.242574836 | 0.009839811 | -0.029167398 | 0.780009899 |

| | | | | | |
|---|---|---|---|---|---|
| Illnesses of father: Parkinson's disease | Both | -0.270393573 | 0.004522213 | 0.330421492 | 0.00402649 |
| Diagnoses - ICD10: N20.0 Calculus of kidney | Male | NA | NA | -0.503786772 | 0.000430899 |
| Vitamin and mineral supplements | Male | 0.004713217 | 0.879244127 | 0.126889535 | 0.000675662 |
| Sources of admission to hospital (recoded): Usual Place of residence | Male | -0.006891964 | 0.862007533 | -0.140296139 | 0.000729451 |
| Methods of discharge from hospital (recoded): Discharged on clinical advice/consent | Male | -0.009427029 | 0.80809518 | -0.130092621 | 0.001325885 |
| Age high blood pressure diagnosed | Male | -0.008057985 | 0.758183537 | -0.075720567 | 0.001340258 |
| Diagnoses - ICD10: I95.9 Hypotension, unspecified | Male | NA | NA | 0.432338452 | 0.003447078 |
| Diagnoses - ICD10: I25.2 Old myocardial infarction | Male | -0.003629735 | 0.980054755 | 0.240779171 | 0.004881535 |
| Diagnoses - ICD10: R69 Unknown and unspecified causes of morbidity | Male | -0.110384936 | 0.131591628 | 0.224040866 | 0.007366213 |
| Glucose | Male | 0.013702507 | 0.377588451 | -0.045358765 | 0.007599488 |
| Mean corpuscular haemoglobin concentration | Male | -0.00224893 | 0.879898208 | 0.042688652 | 0.008526511 |
| Illnesses of father: Stroke | Male | -0.003953195 | 0.924900052 | -0.119250341 | 0.009358861 |

### Figures

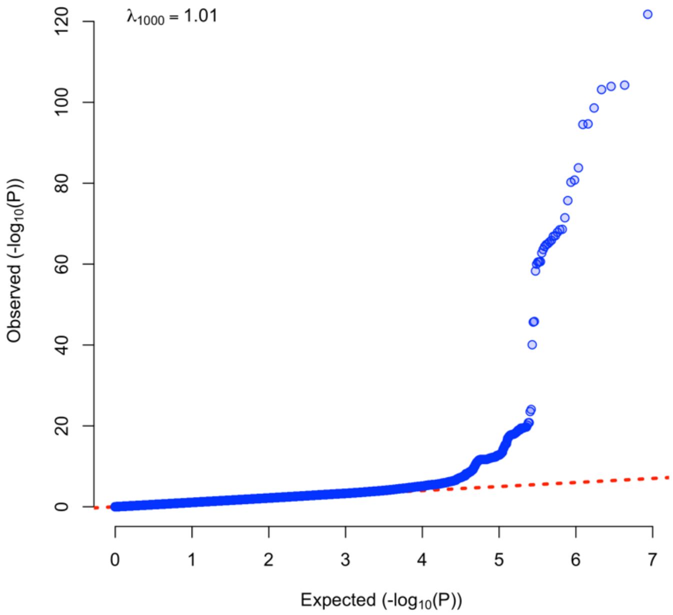

*Supplementary Figure 1: Q-Q Plot of meta-analysis*

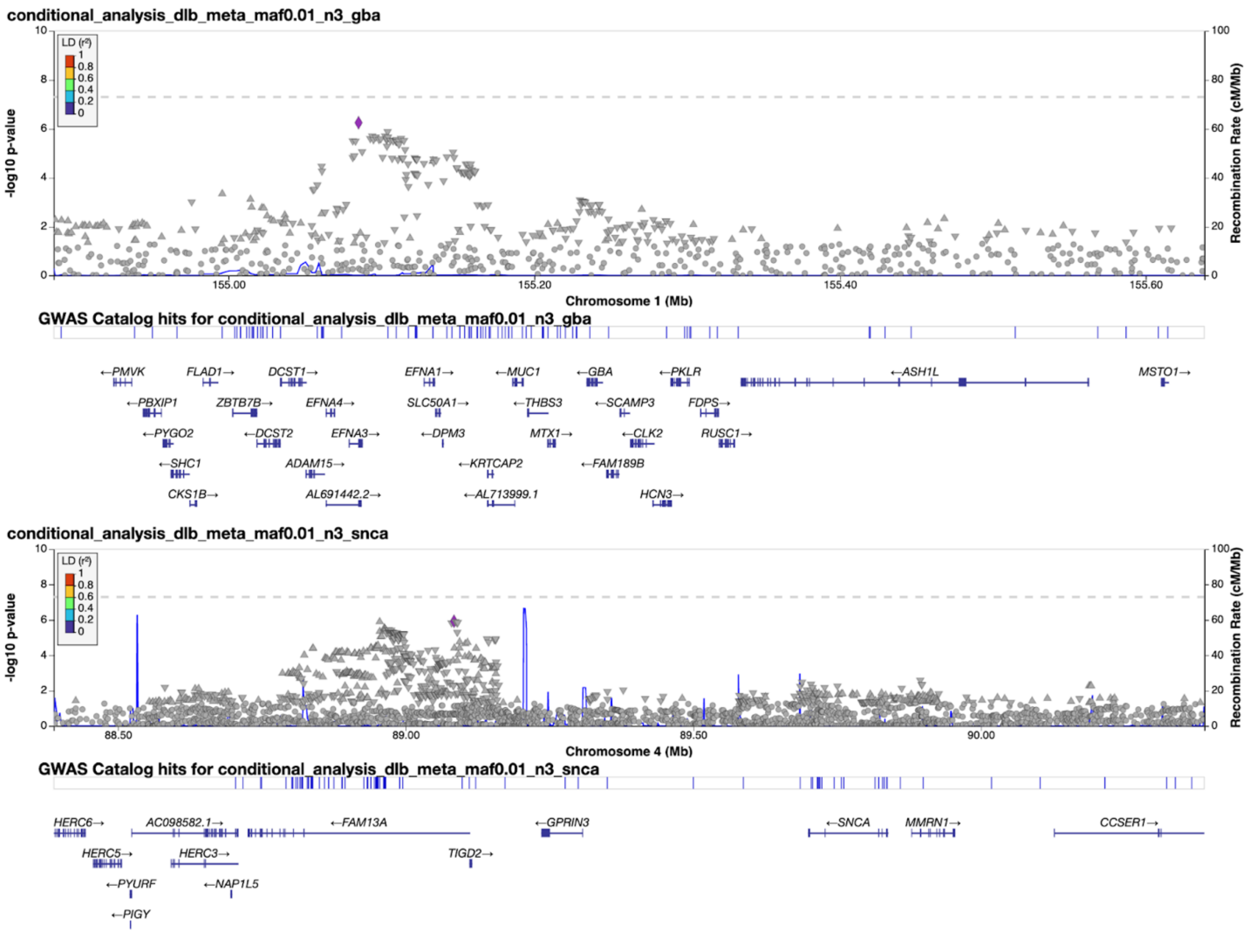

*Supplementary Figure 2: Conditional analysis of SNCA and GBA loci revealed suggestive independent signals.*

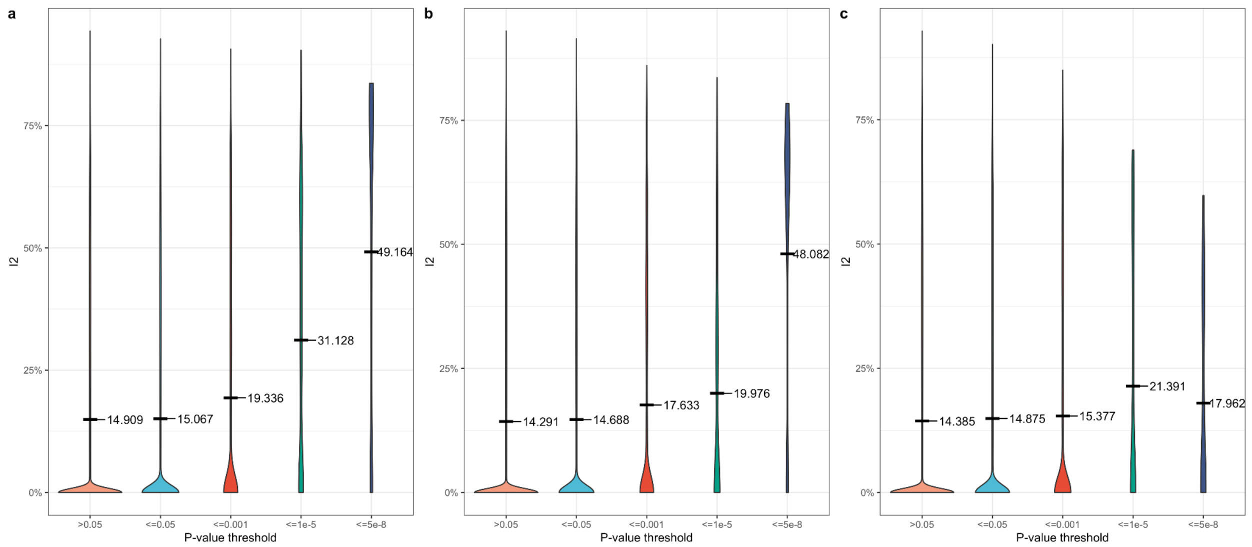

*Supplementary Figure 3: Plots of $I^2$ meta-analysis heterogeneity values for this model. $I^2$ is defined as $I^2 = 100\% \times$ (Cochran's Q-df)/Cochran's Q. Black bar and label denotes group mean. a) Meta-analysis; b) Males from sex-stratified analysis; c) Females from sex-stratified analysis.*

#### The GR@ACE study group

Aguilera Nuria[1], Alarcon Emilio[1], Alegret Montserrat[1,2], Boada Mercè[1,2], Buendia Mar[1], Cano Amanda[1], Cañabate Pilar[1,2], Carracedo Angel[4,5], Corbatón-Anchuelo A[6], de Rojas Itziar[1,2], Diego Susana[1], Espinosa Ana[1,2], Gailhajenet Anna[1], García-González Pablo[1,2], Guitart Marina[1], González-Pérez Antonio[7], Ibarria Marta[1], Lafuente Asunción[1], Macias Juan[8], Maroñas Olalla[4], Martín Elvira[1], Martínez Maria Teresa[6], Marquié Marta[1,2], Montrreal Laura[1], Moreno-Grau Sonia[1,2], Moreno Mariona[1], Raúl Nuñez-Llaves[1], Olivé Clàudia[1] , Orellana Adelina[1], Ortega Gemma[1,2], Pancho Ana[1], Pelejà Ester[1], Pérez-Cordon Alba[1], Pineda Juan A[8], Puerta Raquel[1], Preckler Silvia[1], Quintela Inés[3], Real Luis Miguel[3,8], Rosende-Roca Maitee[1], Ruiz Agustín[1,2], Sáez Maria Eugenia[7], Sanabria Angela[1,2], Serrano-Rios Manuel[6], Sotolongo-Grau Oscar[1], Tárraga Luís[1,2], Valero Sergi[1,2], Vargas Liliana[1].

[1]Research Center and Memory clinic. ACE Alzheimer Center Barcelona, Universitat Internacional de Catalunya, Spain. [2]CIBERNED, Center for Networked Biomedical Research on Neurodegenerative Diseases, National Institute of Health Carlos III, Ministry of Economy and Competitiveness, Spain, [3]Dep. of Surgery, Biochemistry and Molecular Biology, School of Medicine. University of Málaga. Málaga, Spain, [4]Grupo de Medicina Xenómica, Centro Nacional de Genotipado (CEGEN-PRB3-ISCIII). Universidad de Santiago de Compostela, Santiago de Compostela, Spain. [5]Fundación Pública Galega de Medicina Xenómica- CIBERER-IDIS, Santiago de Compostela, Spain. [6]Centro de Investigación Biomédica en Red de Diabetes y Enfermedades Metabólicas Asociadas, CIBERDEM, Spain, Hospital Clínico San Carlos, Madrid, Spain. [7]CAEBI. Centro Andaluz de Estudios Bioinformáticos, Sevilla, Spain, [8]Unidad Clínica de Enfermedades Infecciosas y Microbiología. Hospital Universitario de Valme, Sevilla, Spain.

#### DEGESCO consortium

Adarmes-Gómez Astrid Daniela[1,2], Aguilar Miquel[3,4], Aguilera Nuria[5], Alarcón-Martín Emilio[5], Alcolea Daniel[6,2], Alegret Montserrat[5,2], Alonso María Dolores[7], Alvarez Ignacio[3,4], Álvarez Victoria[8,9], Amer-Ferrer Guillermo[10], Antequera Martirio[11], Antonell Anna[12], Antúnez Carmen[13], Arias Pastor Alfonso[14,15], Baquero Miquel[16], Belbin Olivia[6,2], Bernal Sánchez-Arjona María[17], Boada Mercè[5,2], Buendia Mar[5], Buiza-Rueda Dolores[1,2], Bullido María Jesús[18,2,19,20], Buongiorno Mariateresa[3,4], Burguera Juan Andrés[21], Calero Miguel[22,2,23], Cano Amanda[5], Cañabate Pilar[5], Cardona Serrate Fernando[24,2,25], Carracedo Ángel[26,27], Carrillo Fátima[1,2], Casajeros María José[28], Clarimon Jordi[6,2], Corbatón-Anchuelo Arturo[29,30], Corma-Gómez Anaïs[31], De la Guía Paz[32], de Rojas Itziar[5,2], del Ser Teodoro[33], Diego Susana[5], Diez-Fairen Mónica[3,4], Dols-Icardo Oriol[6,2], Espinosa Ana[5,2], Fernández-Fuertes Marta[31], Fortea Juan[6,2], Franco-Macías Emilio[17,2], Frank-García Ana[34,2,35,20], Gailhajenet Anna[5], García-Alberca Jose María[32], García-González Pablo[5,2], García-Madrona Sebastián[28,36], Garcia-Ribas Guillermo[28,36], Garrote-Espina Lorena[1,2], Gómez-Garre Pilar[1,2], González-Pérez Antonio[37], Guitart Marina[5], Huerto Vilas Raquel[14,15], Ibarria Marta[5], Jesús Silvia[1,2], Labrador Espinosa Miguel Angel[1,2], Lafuente Asunción[5], Lage Carmen[38,2], Legaz Agustina[11], Lleó Alberto[6,2], López-García Sara[39,2], Lopez de Munain Adolfo[40,41,2,42], Macías Juan[31], Macias-García Daniel[1,2], Manzanares Salvadora[11], Marín Marta[17], Marín-Muñoz Juan[11], Maroñas Olalla[26], Marquié Marta[5,2], Martín Elvira[5], Martín Montés Angel[43,2,35], Martínez Begoña[11], Martínez Victoriana[11], Martínez-Lage Álvarez Pablo[44], Martínez-Larrad María Teresa[29,30], Martinez de Pancorbo Marian[45], Martínez Rodríguez

Carmen[46,9], Medina Miguel[2,22], Mendioroz Iriarte Maite[47], Mendoza Silvia[32], Menéndez-González Manuel[48,9,49], Mir Pablo[1,2,50], Molina-Porcel Laura[51,12], Montrreal Laura[5], Moreno Mariona[5], Moreno Fermin[40,2,42], Muñoz-Delgado Laura[1,2], Noguera Perea Fuensanta[11], Núñez-Llaves Raúl[5], Olivé Clàudia[5], Ortega Gemma[5,2], Pancho Ana[5], Pastor Ana Belén[22,52], Pastor Pau[53,54], Pelejá Ester[5], Pérez-Cordón Alba[5], Pérez-Tur Jordi[24,2,25], Periñán María Teresa[1,2], Pineda Juan Antonio[31], Pineda-Sánchez Rocío[1,2], Piñol-Ripoll Gerard[14,15], Preckler Silvia[5], Puerta Raquel[5], Quintela Inés[26], Rábano Alberto[22,52,2], Real Luis Miguel[31,55], Real de Asúa Diego[56], Rodriguez-Rodriguez Eloy[38,2], Rosas Allende Irene[8,9], Rosende-Roca Maitée[5,2], Royo Jose Luís[57], Ruiz Agustín[5,2], Sáez María Eugenia[37], Sanabria Ángela[5,2], Sánchez-Juan Pascual[58,2,59], Sánchez-Valle Raquel[12], Sastre Isabel[18,2], Serrano-Ríos Manuel[29,30], Sotolongo-Grau Oscar[5], Tárraga Lluís[5,2], Valero Sergi[5,2], Vargas Liliana[5], Vicente María Pilar[11], Vivancos-Moreau Laura[11], Zulaica Miren[42,2].

[1]Unidad de Trastornos del Movimiento, Servicio de Neurología y Neurofisiología. Instituto de Biomedicina de Sevilla (IBiS), Hospital Universitario Virgen del Rocío/CSIC/Universidad de Sevilla, Seville, Spain, [2]CIBERNED, Network Center for Biomedical Research in Neurodegenerative Diseases, National Institute of Health Carlos III, Madrid, Spain, [3]Fundació Docència i Recerca MútuaTerrassa, Terrassa, Barcelona, Spain, [4]Memory Disorders Unit, Department of Neurology, Hospital Universitari Mutua de Terrassa, Terrassa, Barcelona, Spain, [5]Research Center and Memory Clinic. Ace Alzheimer Center Barcelona – Universitat Internacional de Catalunya, Spain., [6]Department of Neurology, II B Sant Pau, Hospital de la Santa Creu i Sant Pau, Universitat Autònoma de Barcelona, Barcelona, Spain., [7]Servei de Neurologia. Hospital Clínic Universitari de València, [8]Laboratorio de Genética. Hospital Universitario Central de Asturias, Oviedo, Spain, [9]Instituto de Investigación Sanitaria del Principado de Asturias (ISPA), [10]Department of Neurology, Hospital Universitario Son Espases, Palma, Spain, [11]Unidad de Demencias. Hospital Clínico Universitario Virgen de la Arrixaca, Palma, Spain, [12]Alzheimer's disease and other cognitive disorders unit. Service of Neurology. Hospital Clínic of Barcelona. Institut d'Investigacions Biomèdiques August Pi i Sunyer, University of Barcelona, Barcelona, Spain, [13]Unidad de Demencias, Hospital Clínico Universitario Virgen de la Arrixaca, Murcia, Spain., [14]Unitat Trastorns Cognitius, Hospital Universitari Santa Maria de Lleida, Lleida, Spain, [15]Institut de Recerca Biomedica de Lleida (IRBLLeida), Lleida, Spain, [16]Servei de Neurologia, Hospital Universitari i Politècnic La Fe, Valencia, Spain., [17]Unidad de Demencias, Servicio de Neurología y Neurofisiología. Instituto de Biomedicina de Sevilla (IBiS), Hospital Universitario Virgen del Rocío/CSIC/Universidad de Sevilla, Seville, Spain, [18]Centro de Biología Molecular Severo Ochoa (UAM-CSIC), [19]Instituto de Investigacion Sanitaria 'Hospital la Paz' (IdIPaz), Madrid, Spain, [20]Universidad Autónoma de Madrid, [21]Servei de Neurologia, Hospital Universitari i Politècnic La Fe, Valencia, Spain, [22]CIEN Foundation/Queen Sofia Foundation Alzheimer Center, [23]UFIEC, Instituto de Salud Carlos III, [24]Unitat de Genètica Molecular, Institut de Biomedicina de València-CSIC, Valencia, Spain, [25]Unidad Mixta de Neurologia Genètica, Instituto de Investigación Sanitaria La Fe, Valencia, Spain., [26]Grupo de Medicina Xenómica, Centro Nacional de Genotipado (CEGEN-PRB3-ISCIII). Universidade de Santiago de Compostela, Santiago de Compostela, Spain., [27]Fundación Pública Galega de Medicina Xenómica- CIBERER-IDIS, Santiago de Compostela, Spain., [28]Hospital Universitario Ramon y Cajal, IRYCIS, Madrid, [29]Instituto de Investigación Sanitaria, Hospital Clínico San Carlos (IdISSC), Madrid, Spain, [30]Spanish Biomedical Research Centre in Diabetes and Associated Metabolic Disorders(CIBERDEM), Madrid, Spain, [31]Unidad Clínica de Enfermedades Infecciosas y Microbiología. Hospital Universitario de Valme, Sevilla, Spain, [32]Alzheimer Research Center & Memory Clinic, Instituto Andaluz de Neurociencia, Málaga, Spain., [33]Department of Neurology/CIEN Foundation/Queen Sofia Foundation Alzheimer Center, [34]Department of Neurology, La Paz University Hospital. Instituto de Investigación Sanitaria del Hospital Universitario La Paz. IdiPAZ., [35]Hospital La Paz Institute for Health Research, IdiPAZ, Madrid, Spain, [36]None, [37]CAEBI, Centro Andaluz de Estudios Bioinformáticos, Sevilla, Spain., [38]Neurology Service, Marqués de Valdecilla University Hospital (University of Cantabria and IDIVAL), Santander, Spain., [39]Service of Neurology, University Hospital Marqués de Valdecilla, IDIVAL, University of Cantabria, Santander, Spain, [40]Department of Neurology. Hospital Universitario Donostia. San Sebastian, Spain, [41]Department of Neurosciences. Faculty of Medicine and Nursery. University of the Basque Country, San Sebastián, Spain, [42]Neurosciences Area. Instituto Biodonostia. San Sebastian, Spain, [43]Department of Neurology, La Paz University Hospital, [44]Centro de Investigación y Terapias

Avanzadas. Fundación CITA-alzheimer, San Sebastian, Spain, [45]BIOMICS País Vasco; Centro de investigación Lascaray, Universidad del País Vasco UPV/EHU, Vitoria-Gasteiz, Spain, [46]Hospital de Cabueñes, Gijón, Spain, [47]Navarrabiomed, Pamplona, Spain, [48]Servicio de Neurología. Hospital Universitario Central de Asturias, Oviedo, Spain, [49]Departamento de Medicina, Universidad de Oviedo, Oviedo, Spain, [50]Departamento de Medicina, Facultad de Medicina, Universidad de Sevilla, Seville, Spain., [51]Neurological Tissue Bank of the Biobanc-Hospital Clinic-IDIBAPS, Institut d'Investigacions Biomèdiques August Pi i Sunyer, Barcelona, Spain, [52]BT-CIEN, [53]Unit of Neurodegenerative diseases, Department of Neurology, Hospital Germans Trias i Pujol , Badalona, Barcelona, [54]Neurodegenerative Diseases Research Laboratory, Germans Trias i Pujol Research Laboratory, Badalona, Barcelona, [55]Departamento de Especialidades Quirúrgicas, Bioquímica e Inmunología. Facultad de Medicina. Universidad de Málaga. Málaga, Spain, [56]Hospital Universitario La Princesa, Madrid, Spain, [57]Departamento de Especialidades Quirúrgicas, Bioquímica e Inmunología. School of Medicine. University of Malaga. Málaga, Spain, [58]Alzheimer's Centre Reina Sofia-CIEN Foundation, Centro de Investigación Biomédica en Red sobre Enfermedades Neurodegenerativas (CIBERNED), Madrid, Spain, [59]IDIVAL
